# Prediction of multisite pain incidence in adolescence using a machine learning approach

**DOI:** 10.1101/2023.07.04.23292222

**Authors:** Laura Joensuu, Ilkka Rautiainen, Arto Hautala, Kirsti Siekkinen, Katariina Pirnes, Tuija H Tammelin

**Author notes:** Authors contributed equally (Laura Joensuu, Ilkka Rautiainen). Corresponding Author: Ph.D. Postdoctoral Researcher Laura Joensuu, Faculty of Sport and Health Sciences, University of Jyväskylä, Rautpohjankatu 8, 40700 Jyväskylä, Finland;, @laurajoensuu, Orcid ID: 0000-0002-9544-6552.

## Abstract

**Importance:** Multisite pain is a major adverse health outcome in the adolescent population, affecting the daily lives of up to every third adolescent and their families.

**Objective:** To 1) predict multisite pain incidence in the whole body and in the musculoskeletal locations in adolescents, and 2) explore the sex-specific predictors of multisite pain incidence with a novel machine learning approach.

**Design:** A 2-year observational study (2013-2015). Three different baseline data sets were utilized to predict multisite pain incidence during the follow-up.

**Setting:** Population-based sample of Finnish adolescents.

**Participants:** Apparently healthy adolescents.

**Exposures:** The first data set included 48 selected baseline variables relevant for adolescents’ health and wellbeing. Data included information on students self-reported, objectively measured, and device-based demographics, physical and psychosocial characteristics, and lifestyle factors. The second data set included nine physical fitness variables related to the Finnish national ‘Move!’ monitoring and surveillance system for health-related fitness. The third data set included all available baseline data (392 variables).

**Main Outcome and Measures:** Onset of multisite pain (=weekly pain during the past three months manifesting in at least three sites and not related to any known disease or injury) during the 2-year follow-up in the whole body or musculoskeletal locations. Musculoskeletal pain sites included the neck/shoulder, upper extremities, chest, upper back, low back, buttocks, and lower extremities. Whole body pain sites also the head and abdominal areas. A machine learning algorithm random forest was utilized.

**Results:** Among 410 participants (57% girls) aged on average 12.5-years (SD 1.2), 16 % of boys and 28 % of girls developed multisite pain in the whole body and 10 % and 15 % in the musculoskeletal area during follow-up.

The prediction ability of the machine learning approach with 48 predictive variables reached an AUC 0.65 at highest. With ML, a broad variety of predictors were identified, with up to 33 variables showing predictive power in girls and 13 in boys.

**Conclusions and Relevance:** Findings highlight that rather than any isolated variable, a variety of factors pose a risk for future multisite pain. More emphasis on holistic and multidisciplinary approaches is recommended to prevent multisite pain in adolescence.

**Key points:** 

**Question:** What is the ability of machine learning approach to predict multisite pain incidence during adolescence?

**Findings:** With a random forest machine learning method, a broad variety of predictive physical, lifestyle and psychosocial factors were identified. Prediction ability reached AUC 0.65.

**Meaning:** The findings highlight that predictors of multisite pain incidence in adolescence are multifaceted, although the prediction ability of machine learning remained under clinical relevance (AUC <0.7). These findings support the adoption of holistic and multidisciplinary prevention approaches for multisite pain in adolescence in the future.

## Introduction

Pain is common in adolescents.^1^ Long-lasting pain in at least two bodily locations is reported by at least every tenth and up to every third adolescent in large cohort studies,^2^ with musculoskeletal locations most common sites for pain.^3^ This co-occurrence of pain is typical in the adolescent population,^4^ and hence, multisite pain is recommended to be considered in clinical practice over isolated pain sites.^2^ While the adolescent population is relatively free from many disabling health outcomes, experiences of pain are associated with a lesser ability to conduct daily activities. This association follows a dose-response pattern where more pain sites are associated with a higher degree of disability.^3^ The relevance of multisite pain in adolescence is stressed by pediatric pain researchers.^5^ Pain in adolescence is associated with a broad range of adverse outcomes, from limitations in school attendance and hobbies to reduced quality of life and depressive symptoms.^6–8^ Furthermore, pain experiences tend to track from childhood and adolescence into adulthood,^9, 10^ with relevance to, e.g. future work disability.^11^

Previous studies have found several cross-sectional correlates with pain. Various biological, psychosocial, and lifestyle factors, such as age, pubertal status, overweight or obesity, symptoms of anxiety and depression, chronic health problems, frequent change of residence, poor academic achievement, leisure screen time, fewer interactions with peers, unhealthy lifestyles (e.g. sedentary behavior, screen time, inadequate sleep and smoking), and excessive physical activity, especially in a technical, team, strength, or extreme sports increase the odds for overall or musculoskeletal pain.^1, 2, 12–15^ In addition, physical fitness is suggested to be associated with pain in adolescents. Especially the associations of flexibility and muscular fitness with musculoskeletal pain have been of great interest. Findings remain persistently inconclusive, although the majority of studies have focused on examining a specific pain site.^16, 17^ Despite the inconclusive evidence, proper functioning of the musculoskeletal system, i.e adequate exertion of force, fatigue resistance, and range of motion in the body, is a rationale for many health-related large-scale monitoring and surveillance systems to implement fitness testing at the population level.^16, 18^ Previous findings also indicate that girls report pain more often than boys,^2, 3^ and the correlates of pain might be sex-specific, potentially due to differences in maturation, pain tolerance, or coping behaviors between sexes.^2, 12^

Less is known about the predictors of pain incidence. Predicting the future onset of multisite pain has proven to be challenging despite the broad range of potential explanatory factors. For example, Paananen et al. (2010) did not find statistically significant predictors for multisite musculoskeletal pain incidence in a 2-year follow-up study.^13^ Recently, machine learning (ML)-based pattern recognition approaches have emerged as promising alternatives to traditional statistical approaches in modeling complex phenomenon in various areas of society, including health care.^19^ Therefore, the aim of this study was to 1) predict multisite pain incidence in the musculoskeletal and whole body sites in adolescents and 2) explore the sex-specific determinants of multisite pain incidence utilizing a novel machine learning approach.

## Methods

### Study population

This study was part of a research entity related to the Finnish Schools’ on the Move program focusing on physical activity and wellbeing among children and adolescents.^20^ A longitudinal observational study was conducted between January 2013 and June 2015. A total of 1778 students from nine Finnish schools were invited to participate. Out of these, 970 students (53% girls) provided signed written consent with their main caregiver and participated in the study. After excluding students with possible confounding factors at baseline (such as guardian-reported existing chronic illnesses or disorders, injuries, existing multisite pain, and more than 50% of missing data), the final sample consisted of 410 apparently healthy participants (57% girls) (Figure 1).

**Figure 1.**
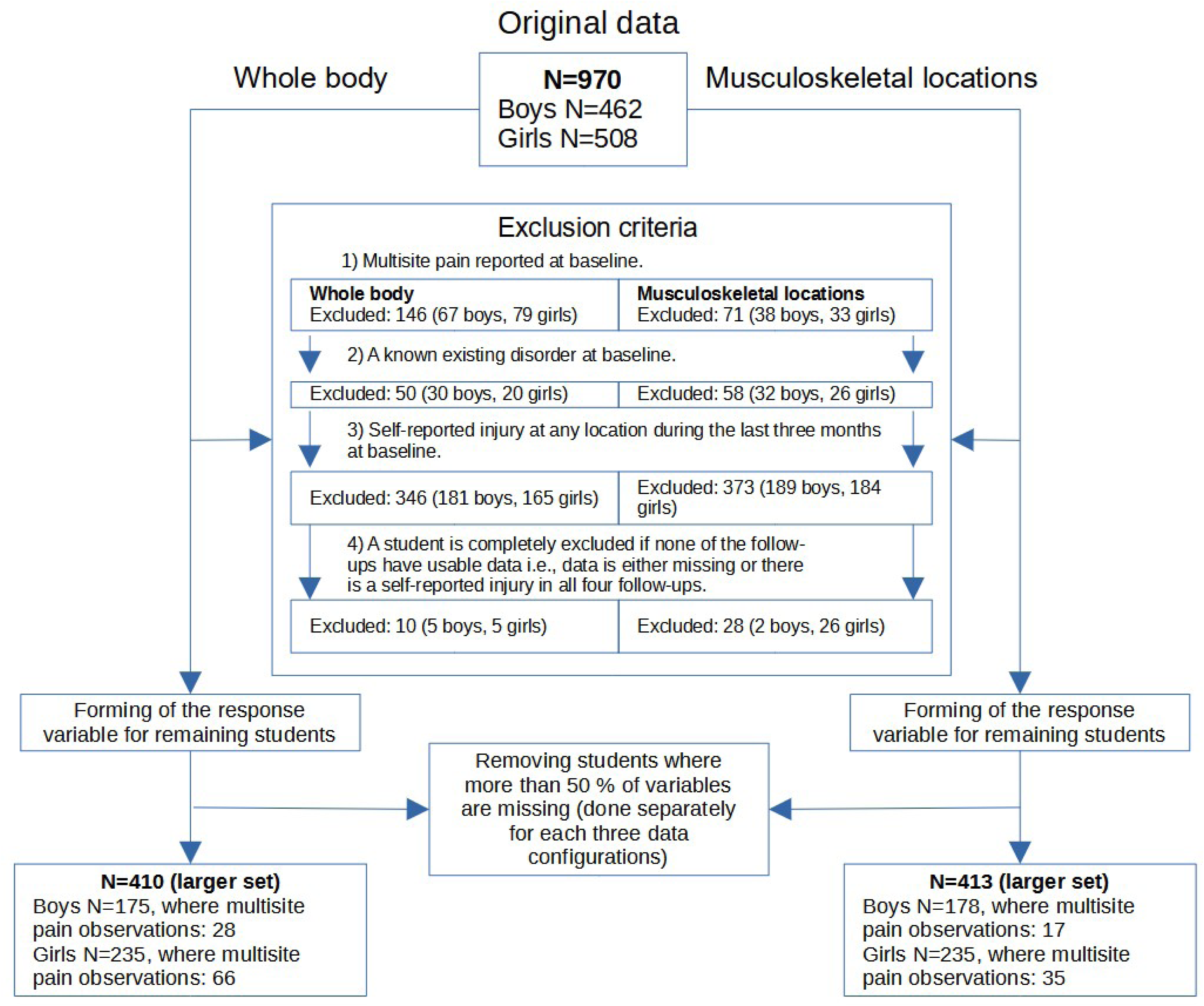
Flow chart of the exclusion process

The study setting and measurements were approved by the Ethics Committee of the University of Jyväskylä, and all procedures were conducted in accordance with the principles outlined in the Declaration of Helsinki. Participants had the option to discontinue their involvement at any point during the research. All measurements were conducted by trained personnel.

### Outcome

Pain incidence was considered the new onset of multisite pain at any time point during the 2-year follow-up. Pain symptoms were screened four times after baseline with a structured questionnaire at six-month intervals: “How often you have had symptoms in the last three months (in body parts A-I in the pictures below)? Mark the appropriate option. Headache (A), Neck and shoulder pain / ache (B), Upper extremities pain / ache (C), Chest pain / ache (D), Upper back pain / ache (E), Low back pain / ache (F), Stomach ache (G), Buttocks pain / ache (H), Lower extremities pain o/ ache (I)”.^21^ The question was supported by an illustration of the described body parts. The answering options were: “Almost daily, More than once a week, About once a week, About once a month, Seldom or never.” Students also reported if the pain originated from trauma: ”Have you injured any of the above-mentioned and pictured pain areas during the previous 3 months (for example, fallen, stumbled, breached during sport, etc.)?” The answer options were “yes” or “no” and provided additional information related to the injured body area.

Multisite pain was defined as reported weekly pain (almost daily, more than once a week, or about once a week) in at least three sites during the past three months. Pains due to traumatic causes were excluded from the analysis. Multisite pain was reported separately for the whole body and musculoskeletal locations. The pain reported in at least three sites was selected to reflect the disabling form of multisite pain.^3^ Musculoskeletal pain sites included the neck/shoulder, upper extremities, chest, upper back, low back, buttocks, and lower extremities. Whole body pain sites included additionally head and abdominal areas.

### Predictive variables

We aimed to determine the predictors of multisite pain incidence using three different data sets. First, we included 48 selected baseline variables relevant to adolescents’ physical activity, fitness, health, and wellbeing.^22^ The data included information on participants’ basic demographics, physical, psychosocial, and lifestyle characteristics and is presented in detail elsewhere.^22^ The results of these analyses are presented in the main text.

Secondly, we used baseline physical fitness measurements belonging to the Finnish national ‘Move!’ monitoring and surveillance system for health-related fitness^23^ (nine baseline variables described in detail elsewhere)^24^. Annually, approximately 100,000 children and adolescents (approximately 96% of the relevant age groups) participate in ‘Move!’ creating a unique database for health-enhancing policies.^18^

Thirdly, a data-driven approach was used with the whole available data (392 baseline variables) to explore potential novel predictors of multisite pain incidence. Data included extensive information on students’ self-reported, objectively measured, and device-based demographics, physical and psychosocial characteristics, and physical activity.

### Analytical procedures

The random forest (RF) method was applied. All analyses were performed using MATLAB R2022b with the Statistics and Machine Learning Toolbox and conducted separately for both sexes. Initial preprocessing and creation of the outcome variable were made using the Python programming language.

RF is an ML method where multiple de-correlated decision trees are grown to form a forest. Afterward, this forest is employed as a voting ensemble, where each tree provides an answer for the prediction task. The final prediction of the forest is the class that gets the most votes from the individual trees.^25, 26^

10-fold cross-validation (CV) was employed for model assessment. During CV, the data for each prediction task was divided into 10 subsamples called folds. Nine of these folds, 90% of the whole data set, were used as the training data to fit the RF model, while one fold, 10% of the data, was used as the validation data. This procedure was repeated ten times in a rotating manner, where eventually all the folds had been employed for training and validation. Thus, all the presented results are based on ten separate data-driven prediction models.

For each of the 10 CV folds, the trained model was employed to predict the out-of-bag (OOB) observations i.e., those observations which were not utilized during the training of each tree, and the validation portion of the data. The main metric recorded was the area the under receiver operating characteristic curve (AUC). T-tests were performed in MATLAB for the OOB and validation data AUC results to determine if the means of the CV folds were significantly (p<0.05) above the random level of 0.5. Further analyses regarding the predictive power of each variable were conducted only in those cases where AUC 95% Confidence Interval did not violate the 0.5 threshold.

RF requires choosing several hyperparameters i.e., options that define the model creation. F-measure for the OOB observations was used as a target during Bayesian optimization,^27^ where several hyperparameters of the RF model were chosen in an automated fashion. Please see Supplementary methods in Supplement 1 for further information on the target measure, the hyperparameters, and other details concerning the RF model.

The contribution of each variable to prediction was estimated using the OOB observations by a permutation importance measure. A baseline result for the model in each CV fold was the accuracy of the model with the original data. To estimate the contribution of each variable, the values of the variables were permuted randomly. The procedure was repeated for all the variables separately, and the accuracy of the model with permutations was recorded for each variable. The accuracy obtained using the permuted variable was then subtracted from the baseline accuracy. The final permutation importance estimate for each variable was the mean of accuracy change for the 10 CV folds. T-tests were employed also for the importance estimates. If the change was significantly (p<0.05) over zero, the variable was seen as having predictive power. Furthermore, if the mean change was near zero or negative, the variable did not have importance in the prediction. MATLAB’s predict function in the TreeBagger class was utilized to calculate the OOB predictions on the trained model. This function computed the weighted average of the class posterior probabilities over the trees.

In further sensitivity analyses the class imbalance was considered, meaning that there are considerably less observations in diffuse idiopathic pain class, is a challenge in all explored settings. This issue was approached during the modelling in two separate ways. Firstly, in the RF model by changing the default cost matrix of misclassification. Cost of misclassifying true pain class observations to no pain class (false negative classification) was increased to 2, while the other misclassification (false positive) was left to its default value 1.

Furthermore, as an alternate view, a synthetic minority oversampling technique for nominal and continuous data SMOTE-NC,^28^ was utilized to see if artificially balancing the training data by oversampling the pain class observations provided any performance improvements. Since SMOTE-NC, available in Themis library in R, expected that there are no missing values in data, missForest imputation for mixed-type data was utilized before oversampling. As a limitation, due to artificially manipulating the training data, the OOB observations could not be meaningfully utilized during this experiment and only non-manipulated validation data for each CV fold was used when estimating the performance measures.

When estimating the importance of individual variables, the associated risk for each variable was examined with simple ROC analysis while acknowledging how the variables were coded in the data. The analysis was done separately from the RF model for the whole age-adjusted data once without utilizing cross-validation. The analysis was performed only for continuous and ordinal variables. The identified risk variables are presented in permutation importance estimate figures with a red panel.

## Results

At baseline, headache (22.5%, 30.4%) and neck and shoulder pain (13.5%, 18.5%) were the most prevalent pain symptoms among boys and girls, respectively (Table 1). Sixteen percent of boys and 28.1% of girls experienced multisite pain incidents in the whole body area during the 2-year follow-up. Multisite pain incidence in the musculoskeletal area was 9.6% and 15.3% in boys and girls, respectively (Table 2).

**Table 1.** Selected subject demographics at baseline by sex.

| N=410 | Boys | Girls |
| --- | --- | --- |
| **Characteristic** |  |  |
| N | 175 (42.6 %) | 235 (57.3 %) |
| Age (years) | 12.5 (1.2) | 12.5 (1.2) |
| Height (cm) | 156.7 (10.9) | 155.1 (9.6) |
| Weight (kg) | 46.1 (12.2) | 45.4 (10.1) |
| BMI (kg/m<sup>2</sup>) | 18.5 (3.2) | 18.7 (3.0) |
| **Pain at different body sites** |  |  |
| **Musculoskeletal pain sites** |  |  |
| Neck/Shoulder (%) | 23 (13.6 %) | 42 (18.5 %) |
| Upper extremities (%) | 7 (4.1 %) | 10 (4.4 %) |
| Chest (%) | 1 (0.6 %) | 2 (0.9 %) |
| Upper back (%) | 5 (3.0 %) | 5 (2.2 %) |
| Low back (%) | 8 (4.7 %) | 8 (3.5 %) |
| Buttocks (%) | 2 (1.2 %) | 6 (2.6 %) |
| Lower extremities (%) | 9 (5.3 %) | 13 (5.7 %) |
| **Other pain sites** |  |  |
| Head (%) | 38 (22.5 %) | 69 (30.4 %) |
| Abdominal (%) | 18 (10.7 %) | 36 (15.9 %) |
| **Physical activity and fitness** |  |  |
| Moderate-to-vigorous physical activity (min/day) | 58.3 (24.2) | 47.4 (18.3) |
| Sedentary time (hours/day) | 8.1 (1.3) | 8.6 (1.1) |
| Physical fitness index (Move-index) | 16.6 (4.1) | 17.2 (3.6) |

Values are means and standard deviations for continues variables and proportions of participants for others. BMI, Body mass index; Move-index, weighted sum of the Finnish national Move! monitoring system's fitness items.

**Table 2.**
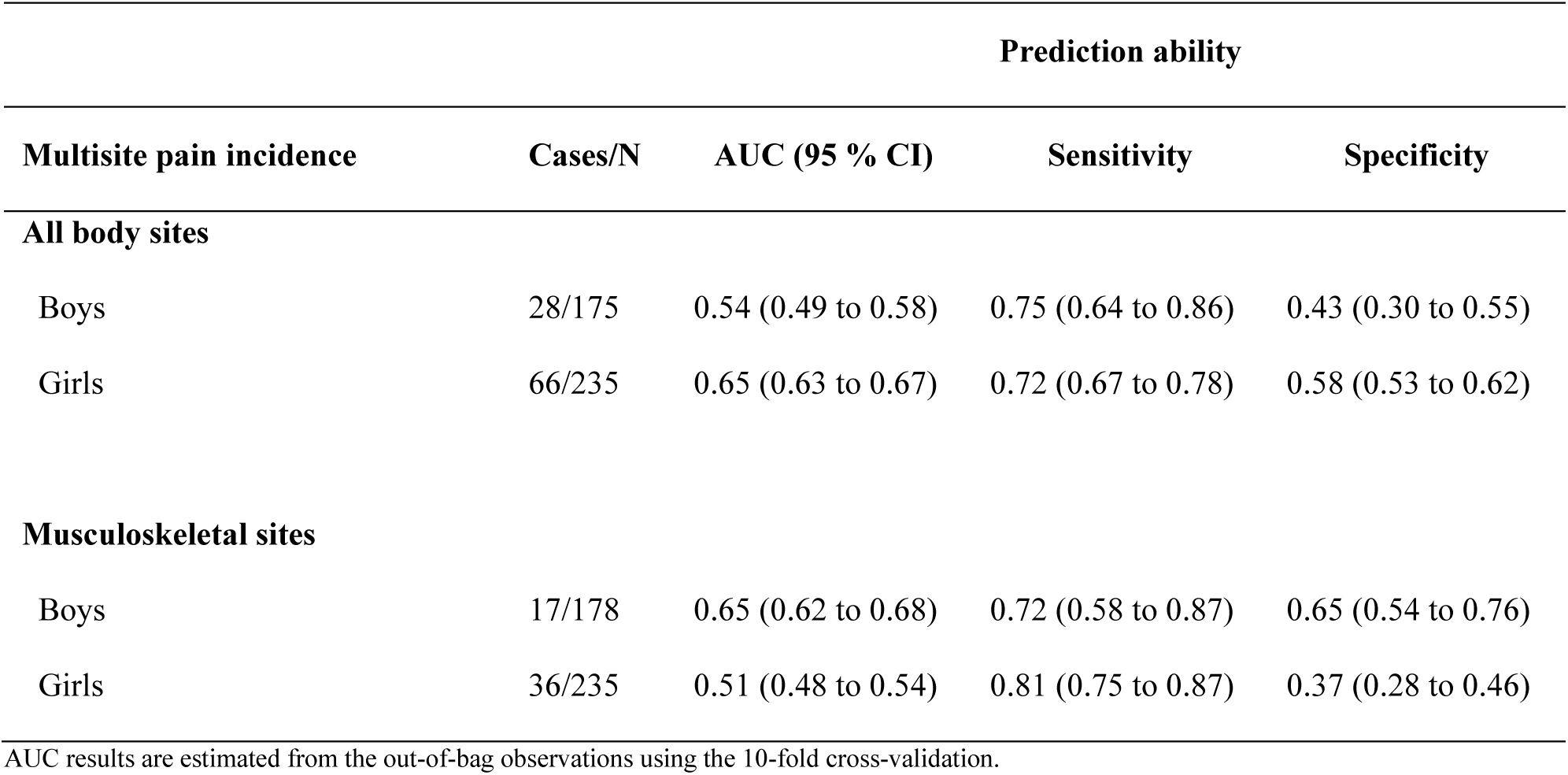
Prediction ability of machine learning for multisite pain incidence among adolescents.

| Prediction ability |  |  |  |  |
| --- | --- | --- | --- | --- |
| Multisite pain incidence | Cases/N | AUC (95 % CI) | Sensitivity | Specificity |
| All body sites |  |  |  |  |
| Boys | 28/175 | 0.54 (0.49 to 0.58) | 0.75 (0.64 to 0.86) | 0.43 (0.30 to 0.55) |
| Girls | 66/235 | 0.65 (0.63 to 0.67) | 0.72 (0.67 to 0.78) | 0.58 (0.53 to 0.62) |
| Musculoskeletal sites |  |  |  |  |
| Boys | 17/178 | 0.65 (0.62 to 0.68) | 0.72 (0.58 to 0.87) | 0.65 (0.54 to 0.76) |
| Girls | 36/235 | 0.51 (0.48 to 0.54) | 0.81 (0.75 to 0.87) | 0.37 (0.28 to 0.46) |

AUC results are estimated from the out-of-bag observations using the 10-fold cross-validation.

The ability of the machine learning approach to predict whole body multisite pain incidence reached an AUC 0.54 (95% Confidence Interval 0.49 to 0.58) for boys and 0.65 (0.63 to 0.67) for girls (Table 2). The prediction ability for multisite musculoskeletal pain incidence was AUC 0.65 (0.62 to 0.68) in boys and 0.51 (0.48 to 0.54) in girls.

The tasks where prediction ability reached above random level (AUC >0.5) were further analyzed for variable importance. Altogether, 33 variables out of 48 baseline variables showed predictive power for whole body multisite pain incidence among girls. All variables are illustrated in Figure 2, and the top ten are described in detail here. Poorer perceived health, higher perceived fitness, more frequent tiredness on schoolday mornings, having overweight or obesity based on body mass index, more frequent participation in sports competitions and matches, more frequent breakfast eating during the school week, a lower grade point in physical education, a lower amount moderate-to-vigorous physical activity during leisure time, higher school enjoyment, and higher pubertal status increased the probability of multisite pain incidence in the whole body area in girls.

**Figure 2.**
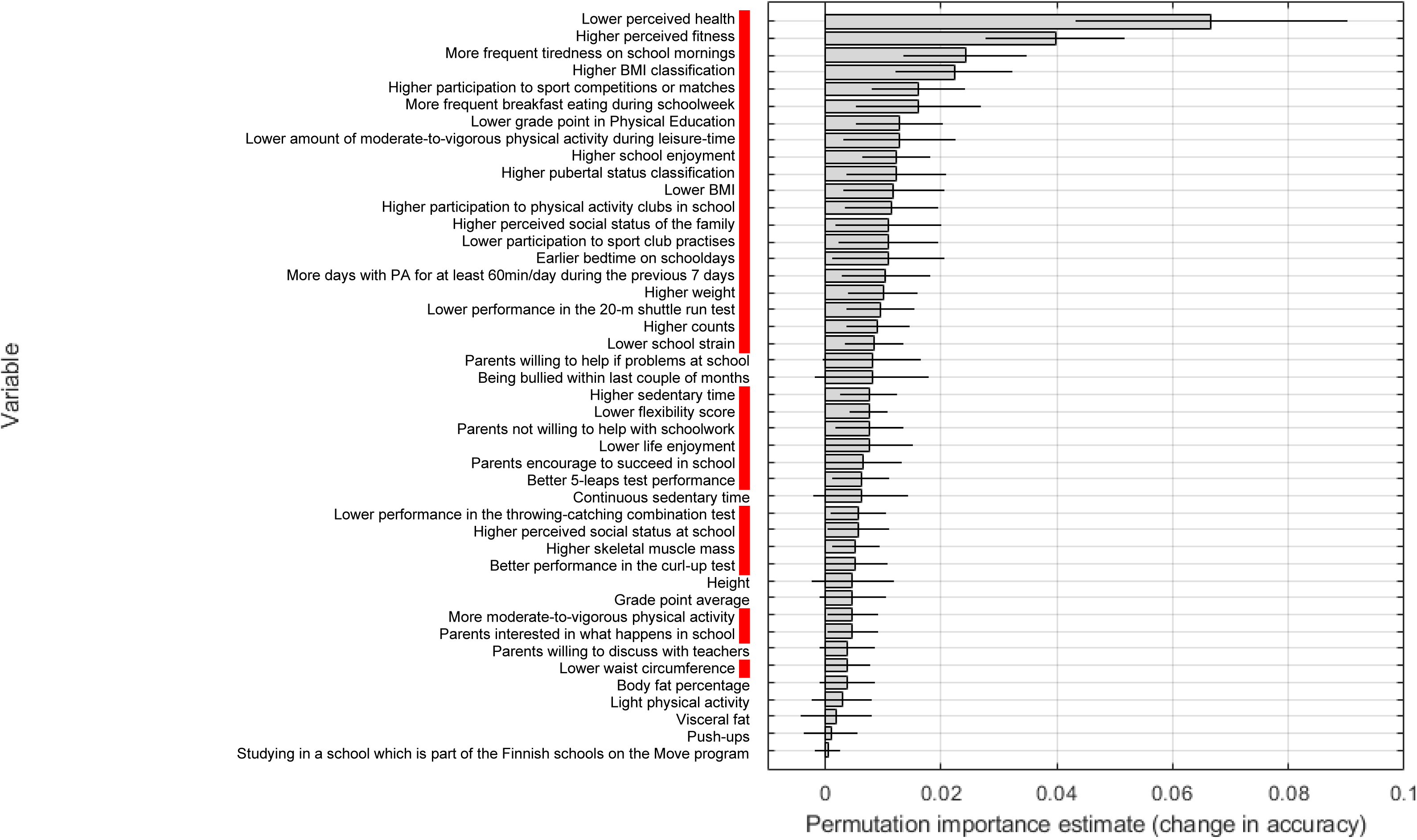
Permutation importance estimates for girls in the *selected **set*** for *all sites* (AUC 0.65). Red panel, risk factors; PA, physical activity; Counts, accelerometer total activity counts

In boys, a total of 13 variables out of 48 showed predictive power for multisite musculoskeletal pain incidence. The top ten predictors for pain incidence included higher school strain, lower school enjoyment, a higher participation rate in sports competitions or matches, lower amounts of continuous device-measured sedentary time, better muscular fitness measured with the number of push-ups conducted within one minute, a lower body mass index, more active participation in physical activity clubs in school, a later bedtime on schooldays, lower total sedentary time, and parents’ higher willingness to help with schoolwork (Figure 3).

**Figure 3.**
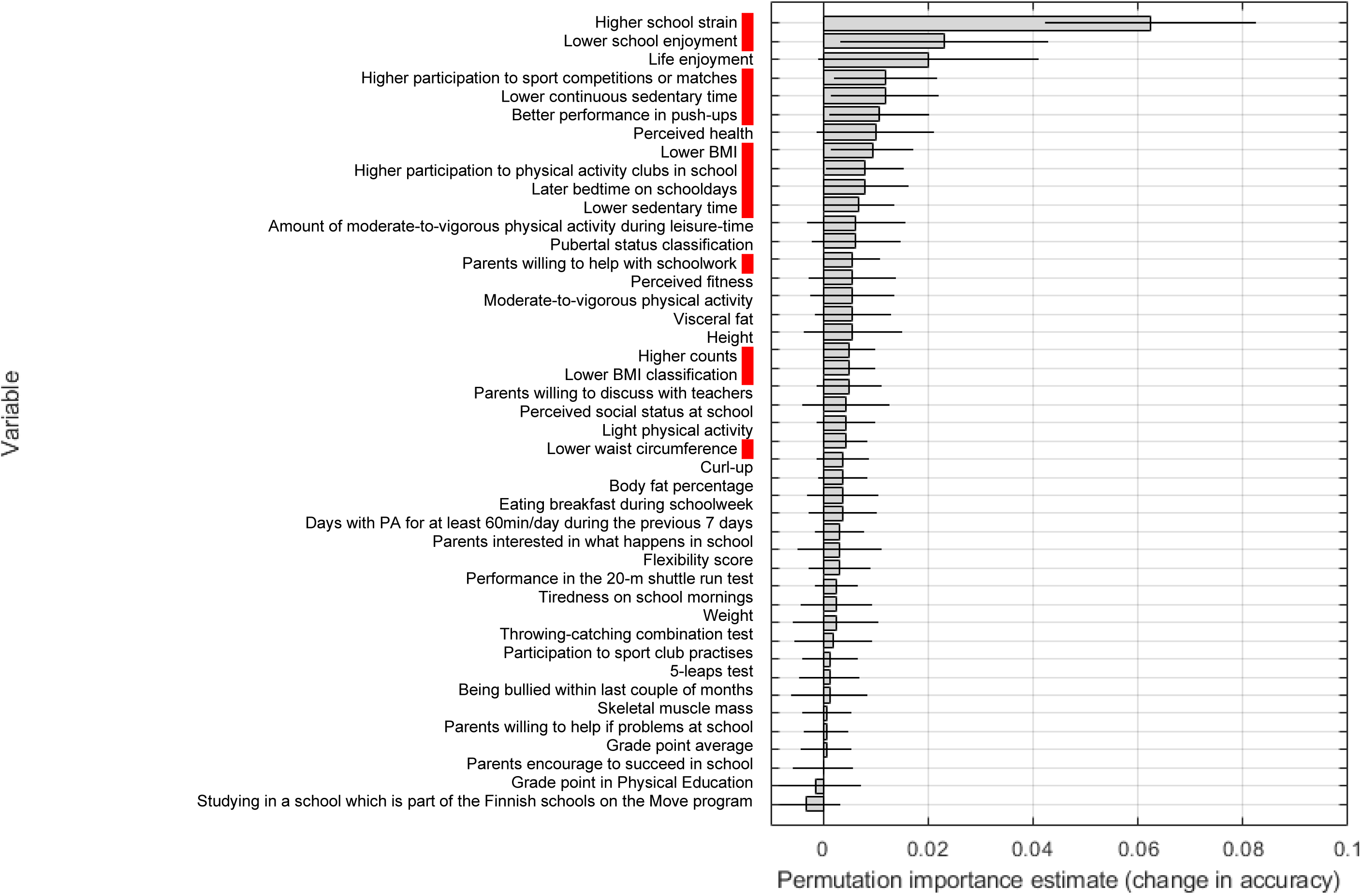
Permutation importance estimates for boys in the *selected set* for *musculoskeletal sites* (AUC 0.65). Red panel, risk factors; PA, physical activity; Counts, accelerometer total activity counts

Prediction ability with the Move! variables reached above the random level only in boys and in the whole body area (AUC 0.59 (0.56 to 0.62), eTable 1) and indicated that better muscular and cardiorespiratory fitness but poorer motor fitness predict higher multisite pain incidence (eFigure 1). With the full available data set ML, was able to predict multisite pain incidence only in girls (AUC 0.68 (0.66 to 0.70) and 0.58 (0.56 to 0.60) in whole body and musculoskeletal sites, respectively) (eTable 2). With the full data, along with physical, psychosocial, and lifestyle factors, individual pain sites at baseline rose as predictive factors of future multisite pain (eFigures 2 and 3). Balancing the data artificially with SMOTE-NC did improve prediction ability (up to AUC 0.72, with high a standard deviation as a result of the small size of each validation fold), but due to automatic risk threshold selection designed for earlier tasks, it created suboptimal sensitivity and specificity values (eTables 3-5).

## Discussion

In this study, we aimed to investigate determinants of multisite pain incidence among the adolescent population with a novel ML approach. Multisite pain incidence in the study population was considerable, with up to 16% of boys and 28% of girls developing multisite weekly pain during the 2-year follow-up. The prediction ability of the ML approach with selected predictive variables reached an AUC 0.65 at its highest. With ML, a broad variety of variables predicting multisite pain incidence in adolescents were identified. Out of 48 selected variables, up to 33 variables showed predictive power in girls and 13 in boys. These findings highlight that rather than any isolated variable, a variety of factors may possess an increased risk for multisite pain and indicate the paradoxical nature of some variables, especially in girls.

Multisite pain is a major adverse health outcome in the adolescent population, affecting the daily lives of more than every fourth adolescent and their families.^1^ Predicting the future onset of multisite pain, identifying individuals potentially experiencing disabling pain in the future, and recognizing the predictors of pain hold the potential to enhance the quality of life in this important demographic through better health education and policies. Pediatric experts have long stressed the importance of pain research and further understanding of pain epidemiology and underlying pathophysiology through innovative study designs.^5^

Machine learning-based pattern recognition algorithms, a subgroup of artificial intelligence, have emerged as promising alternatives to traditional statistical methods in developing next-generation tools to enhance public health. In contrast to theory-based and often restricted traditional statistical models, the ML approach enables near unlimited learning capacity from the available data,^25^ providing the potential to develop more precise methods for screening and predicting adverse health outcomes. ML-based approaches are acknowledged to hold significant potential for reforming public health policies in the future.^29^

Previous studies have shown that prediction of multisite pain incidence is demanding,^13^ and the isolated correlates have modest effect sizes.^8^ In this current study, the ML approach was able to predict pain incidence above the random level, however remaining under clinical relevance (AUC <0.7).^30^ Through the ML approach, we found various predictors for multisite pain incidence, reflecting the previously reported physical, lifestyle, and psychosocial correlates,^1, 2, 12–15^ and complementing these findings by illustrating the risky variables in a holistic framework alongside the paradoxical nature of some variables. For example, with whole body multisite pain incidence among girls, indicators of both lower and higher psychosocial wellbeing (e.g. lower life enjoyment vs. higher school enjoyment), low and high physical activity (lower amount of moderate-to-vigorous physical activity during leisure time vs. more days with physical activity for at least 60 min per day), lower body mass index, and obesity or overweight classification were identified as risk factors. These findings illustrate that risk factors, especially for whole body multisite pain incidence in girls are complex, associations are not linear, and individuals with both healthy and unhealthy lifestyles, favorable or unfavorable psychosocial status might develop multisite pain in the future. In boys, findings indicated more consistently that poorer psychosocial wellbeing, higher physical activity, a leaner body, and better physical fitness predict multisite musculoskeletal pain incidence and support the acknowledgment of overall wellbeing and health-enhancing physical activity practices among boys to prevent musculoskeletal pain.

The strengths of this study were the novel application of ML in pain prediction, the longitudinal study design, and the extensiveness of predictors. The ML approach considerably extends pain research and provides potential avenues for screening and modeling complex phenomena in the future. The data was however limited by information (no data on current medication) and cases (e.g., <66 cases in the data set) with possibly influencing the generalizability of the findings. ML explores patterns in the data and does not explain underlying mechanisms or causality. The multicollinearity of the variables might affect the interpretation of variables with similar phenomenal origins. Self-reported data may suffer from recall bias, although the reliability of the utilized questionnaire has shown to be reasonable.^21^

In conclusion, these novel findings highlight the multifaceted predictors of multisite pain incidence in adolescents and support the adoption of holistic and multidisciplinary prevention approaches in the future.

## Statements

This study was funded by Ella and Georg Ehrnrooth foundation. Data collection for this study was supported by the Juho Vainio Foundation (201410342) and the Finnish Ministry of Education and Culture (OKM/92/626/2013).

## Data sharing

Data and utilized scripts are available upon reasonable request from IR (scripts) and THT (data).

## Declaration of interests

We declare no competing interests

## Data Availability

Data and utilized scripts are available upon reasonable request from IR (scripts) and THT (data).

## Notes

### Competing Interest Statement

The authors have declared no competing interest.

### Author Declarations

The study setting and measurements were approved by the Ethics Committee of the University of Jyvaskyla

## References

1. King S, Chambers CT, Huguet A, et al. The epidemiology of chronic pain in children and adolescents revisited: A systematic review. Pain. 2011;152(12):2729–2738. doi:10.1016/j.pain.2011.07.016

2. Gobina I, Villberg J, Välimaa R, et al. Prevalence of self-reported chronic pain among adolescents: Evidence from 42 countries and regions. Eur J Pain. 2019;23(2):316–326. doi:10.1002/ejp.1306

3. Hoftun GB, Romundstad PR, Zwart JA, Rygg M. Chronic idiopathic pain in adolescence – high prevalence and disability: The young HUNT study 2008. Pain. 2011;152(10):2259–2266. doi:10.1016/j.pain.2011.05.007

4. Kujala UM, Taimela S, Viljanen T. Leisure physical activity and various pain symptoms among adolescents. Br J Sports Med. 1999;33(5):325–328. doi:10.1136/bjsm.33.5.325

5. López-Solà M, Suñol M, Timmers I. Brain predictors of multisite pain onset in children. Pain. 2022;163(4):e502–e503. doi:10.1097/j.pain.0000000000002430

6. Holden S, Rathleff MS, Roos EM, Jensen MB, Pourbordbari N, Graven-Nielsen T. Pain patterns during adolescence can be grouped into four pain classes with distinct profiles: A study on a population based cohort of 2953 adolescents. Eur J Pain. 2018;22(4):793–799. doi:10.1002/ejp.1165

7. Gauntlett-Gilbert J, Eccleston C. Disability in adolescents with chronic pain: Patterns and predictors across different domains of functioning. Pain. 2007;131(1):132–141. doi:10.1016/j.pain.2006.12.021

8. Auvinen J, Eskola PJ, Ohtonen HR, et al. Long-term adolescent multi-site musculoskeletal pain is associated with psychological distress and anxiety. J Psychosom Res. 2017;93:28–32. doi:10.1016/j.jpsychores.2016.12.006

9. Lucas R, Brandão M, Gorito V, Talih M. Refining the prediction of multisite pain in 13-year-old boys and girls by using parent-reported pain experiences in the first decade of life. Eur J Pain. 2022;26(3):695–708. doi:10.1002/ejp.1898

10. Kamaleri Y, Natvig B, Ihlebaek CM, Benth JS, Bruusgaard D. Change in the number of musculoskeletal pain sites: A 14-year prospective study. Pain. 2009;141(1):25–30. doi:10.1016/j.pain.2008.09.013

11. Kamaleri Y, Natvig B, Ihlebaek CM, Bruusgaard D. Does the number of musculoskeletal pain sites predict work disability? A 14-year prospective study. Eur J Pain. 2009;13(4):426–430. doi:10.1016/j.ejpain.2008.05.009

12. Hoftun GB, Romundstad PR, Rygg M. Factors Associated With Adolescent Chronic Non-Specific Pain, Chronic Multisite Pain, and Chronic Pain With High Disability: The Young–HUNT Study 2008. J Pain. 2012;13(9):874–883. doi:10.1016/j.jpain.2012.06.001

13. Paananen MV, Taimela SP, Auvinen JP, et al. Risk factors for persistence of multiple musculoskeletal pains in adolescence: A 2-year follow-up study. Eur J Pain. 2010;14(10):1026–1032. doi:10.1016/j.ejpain.2010.03.011

14. Guddal MH, Stensland SØ, Småstuen MC, Johnsen MB, Zwart JA, Storheim K. Physical Activity Level and Sport Participation in Relation to Musculoskeletal Pain in a Population-Based Study of Adolescents: The Young-HUNT Study. Orthop J Sports Med. 2017;5(1):232596711668554. doi:10.1177/2325967116685543

15. Pirnes KP, Kallio J, Hakonen H, Hautala A, Häkkinen AH, Tammelin T. Physical activity, screen time and the incidence of neck and shoulder pain in school-aged children. Sci Rep. 2022;12(1):10635. doi:10.1038/s41598-022-14612-0

16. Plowman S, Meredith M. Fitnessgram/Activitygram ReferenceGuide. 4th ed. The Cooper Institute; 2013.

17. Pirnes KP, Kallio JJ, Hakonen HJ, et al. Physical fitness characteristics and neck and shoulder pain incidence in school-aged children—A 2-year follow-up. Health Sci Rep. 2022;5(6). doi:10.1002/hsr2.852

18. Joensuu L, Csányi T, Huhtiniemi M, et al. How to Design and Establish a National School-Based Physical Fitness Monitoring and Surveillance System for Children and Adolescents: The Ten-Step Approach Recommended by the FitBack Network. Open Science Framework; 2023. doi:10.31219/osf.io/zsnju

19. Gevaert AB, Adams V, Bahls M, et al. Towards a personalised approach in exercise-based cardiovascular rehabilitation: How can translational research help? A ‘call to action’ from the Section on Secondary Prevention and Cardiac Rehabilitation of the European Association of Preventive Cardiology. Eur J Prev Cardiol. 2020;27(13):1369–1385. doi:10.1177/2047487319877716

20. Blom A, Tammelin T, Laine K, Tolonen H. Bright spots, physical activity investments that work: the Finnish Schools on the Move programme. Br J Sports Med. 2018;52(13):820–822. doi:10.1136/bjsports-2017-097711

21. Pirnes KP, Kallio J, Siekkinen K, Hakonen H, Häkkinen A, Tammelin T. Test-retest repeatability of questionnaire for pain symptoms for school children aged 10–15 years. Scand J Pain. 2019;19(3):575–582. doi:10.1515/sjpain-2018-0338

22. Joensuu L, Rautiainen I, Äyrämö S, et al. Precision exercise medicine: predicting unfavourable status and development in the 20-m shuttle run test performance in adolescence with machine learning. BMJ Open Sport Exerc Med. 2021;7(2):e001053. doi:10.1136/bmjsem-2021-001053

23. Move! https://www.oph.fi/en/move

24. Joensuu L, Syväoja H, Kallio J, Kulmala J, Kujala UM, Tammelin TH. Objectively measured physical activity, body composition and physical fitness: Cross-sectional associations in 9-to 15-year-old children. Eur J Sport Sci. 2018;18(6):882–892. doi:10.1080/17461391.2018.1457081

25. Breiman L. Random Forests. Mach Learn. 2001;45(1):5-32. doi:10.1023/A:1010933404324

26. Hastie T, Friedman J, Tibshirani R. The Elements of Statistical Learning. Springer New York; 2001. doi:10.1007/978-0-387-21606-5

27. Snoek J, Larochelle H, Adams RP. Practical Bayesian Optimization of Machine Learning Algorithms. In: Pereira F, Burges CJ, Bottou L, Weinberger KQ, eds. Advances in Neural Information Processing Systems. Vol 25. Curran Associates, Inc.; 2012. https://proceedings.neurips.cc/paper_files/paper/2012/file/05311655a15b75fab86956663e1819cd-Paper.pdf

28. Chawla NV, Bowyer KW, Hall LO, Kegelmeyer WP. SMOTE: Synthetic Minority Over-sampling Technique. J Artif Intell Res. 2002;16:321–357. doi:10.1613/jair.953

29. Horizon Europe Work Programme 2023-2024, 4. Health.; 2022.

30. Hosmer D, Lemeshow S. Chapter 5. In: Applied Logistic Regression. 2nd ed. John Wiley and Sons; 2000:160–164.

