## Supplement 1 for "Prediction of multisite pain incidence in adolescence using a machine learning approach"

*Authors contributed equally

**4 eFigures**

eFigure 1. Flow chart of the exclusion process
eFigure 2. Permutation importance estimates for boys in the *Move! set* for *all sites* (AUC 0.59).
eFigure 3. Permutation importance estimates for girls in the *full set* for *all sites* (AUC 0.68).
eFigure 4. Permutation importance estimates for girls in the *full set* for *musculoskeletal sites* (AUC 0.58).

**5 eTables**

eTable 1. Prediction ability of machine learning for multisite pain incidence among adolescents. Results for the Move! set.
eTable 2. Prediction ability of machine learning for multisite pain incidence among adolescents. Results for the full set.
eTable 3. Prediction ability of machine learning for multisite pain incidence among adolescents. Results for the Move! set, balanced with SMOTE-NC.
eTable 4. Prediction ability of machine learning for multisite pain incidence among adolescents. Results for the selected data set, balanced with SMOTE-NC.
eTable 5. Prediction ability of machine learning for multisite pain incidence among adolescents. Results for the full set, balanced with SMOTE-NC.

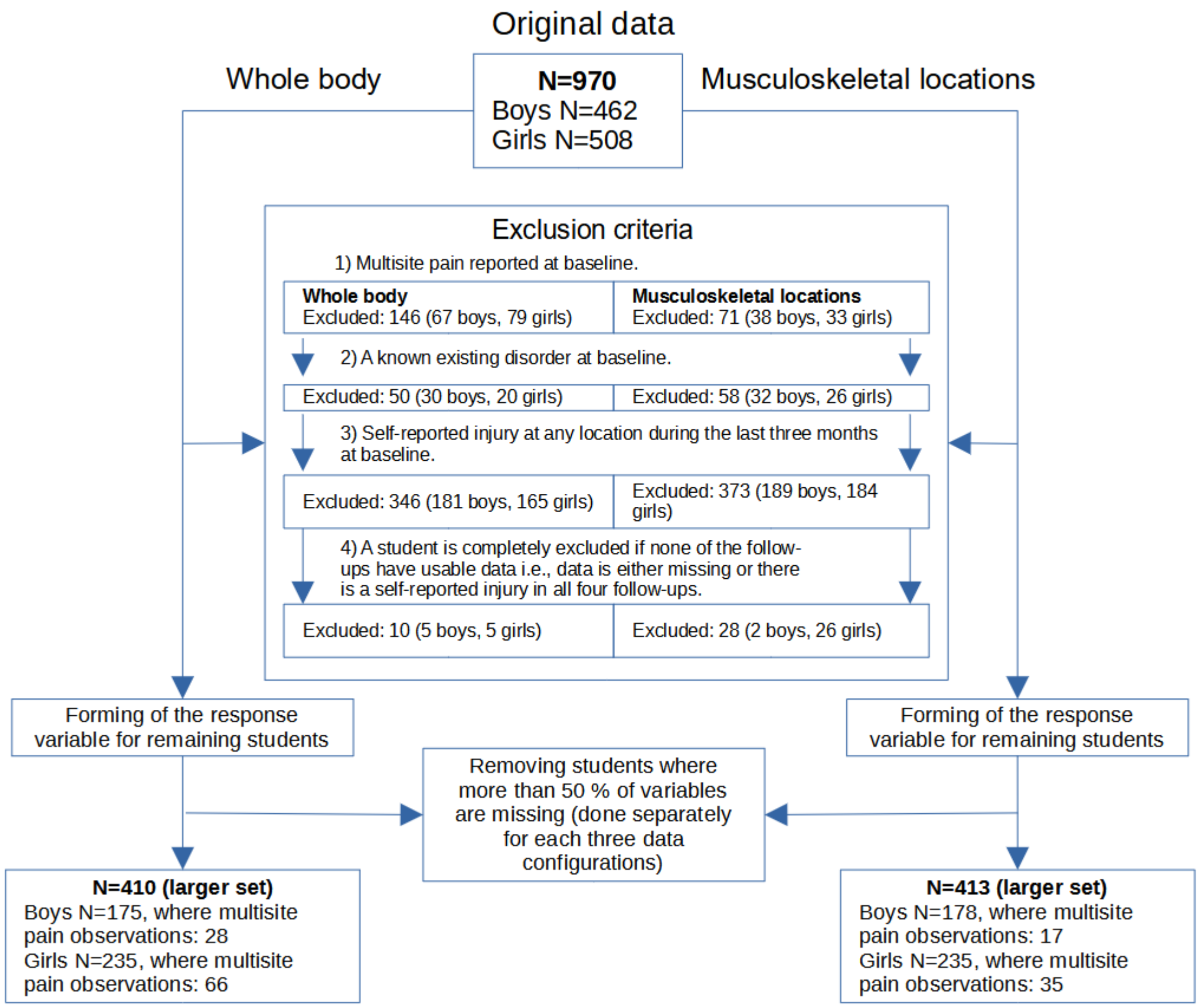


**eFigure 1.** Flow chart of the exclusion process

**eTable 1.** Prediction ability of machine learning for multisite pain incidence among adolescents. Results for the Move! set.

| | | Prediction ability | | |
|---|---|---|---|---|
| **Multisite pain incidence** | **Cases/N** | **AUC (95 % CI)** | **Sensitivity** | **Specificity** |
| **All body sites** | | | | |
| Boys | 25/168 | 0.59 (0.56 to 0.62) | 0.69 (0.60 to 0.78) | 0.59 (0.50 to 0.68) |
| Girls | 63/225 | 0.38 (0.34 to 0.41) | 0.98 (0.98 to 0.98) | 0.01 (0.00 to 0.02) |
| **Musculoskeletal sites** | | | | |
| Boys | 17/170 | 0.44 (0.40 to 0.47) | 0.71 (0.53 to 0.90) | 0.40 (0.18 to 0.63) |
| Girls | 34/225 | 0.35 (0.31 to 0.40) | 0.85 (0.72 to 0.99) | 0.17 (0.00 to 0.34) |

AUC results are estimated from the out-of-bag observations using the 10-fold cross-validation.

**eTable 2.** Prediction ability of machine learning for multisite pain incidence among adolescents. Results for the full set.

| | | Prediction ability | | |
|---|---|---|---|---|
| **Multisite pain incidence** | **Cases/N** | **AUC (95 % CI)** | **Sensitivity** | **Specificity** |
| **All body sites** | | | | |
| Boys | 28/169 | 0.46 (0.42 to 0.50) | 0.89 (0.85 to 0.93) | 0.19 (0.13 to 0.26) |
| Girls | 65/230 | 0.68 (0.66 to 0.70) | 0.64 (0.55 to 0.74) | 0.65 (0.52 to 0.78) |
| **Musculoskeletal sites** | | | | |
| Boys | 18/172 | 0.54 (0.49 to 0.59) | 0.69 (0.56 to 0.82) | 0.51 (0.37 to 0.65) |
| Girls | 35/230 | 0.58 (0.56 to 0.60) | 0.63 (0.53 to 0.73) | 0.61 (0.49 to 0.73) |

AUC results are estimated from the out-of-bag observations using the 10-fold cross-validation.

**eTable 3.** Prediction ability of machine learning for multisite pain incidence among adolescents. Results for the Move! set, balanced with SMOTE-NC.

| | | Prediction ability | | |
|---|---|---|---|---|
| **Multisite pain incidence** | **Cases/N** | **AUC (95 % CI)** | **Sensitivity** | **Specificity** |
| **All body sites** | | | | |
| Boys | 25/168 | 0.65 (0.54 to 0.77) | 0.35 (0.19 to 0.52) | 0.80 (0.72 to 0.88) |
| Girls | 63/225 | 0.44 (0.39 to 0.49) | 0.39 (0.27 to 0.51) | 0.54 (0.46 to 0.63) |
| **Musculoskeletal sites** | | | | |
| Boys | 17/170 | 0.52 (0.37 to 0.66) | 0.15 (0.00 to 0.30) | 0.85 (0.80 to 0.91) |
| Girls | 34/225 | 0.30 (0.21 to 0.40) | 0.17 (0.05 to 0.28) | 0.64 (0.54 to 0.75) |

Results are estimated from the validation set observations using the 10-fold cross-validation.

**eTable 4.** Prediction ability of machine learning for multisite pain incidence among adolescents. Results for the selected data set, balanced with SMOTE-NC.

| | | Prediction ability | | |
|---|---|---|---|---|
| **Multisite pain incidence** | **Cases/N** | **AUC (95 % CI)** | **Sensitivity** | **Specificity** |
| **All body sites** | | | | |
| Boys | 28/175 | 0.61 (0.52 to 0.71) | 0.00[a] | 0.98 (0.95 to 1.00) |
| Girls | 66/235 | 0.44 (0.39 to 0.49) | 0.39 (0.27 to 0.51) | 0.54 (0.46 to 0.63) |
| **Musculoskeletal sites** | | | | |
| Boys | 17/178 | 0.72 (0.61 to 0.82) | 0.65 (0.40 to 0.91) | 0.63 (0.50 to 0.76) |
| Girls | 36/235 | 0.50 (0.42 to 0.58) | 0.03 (0.00 to 0.10) | 0.95 (0.92 to 0.98) |

Results are estimated from the validation set observations using the 10-fold cross-validation. [a]Due to automated threshold selection that maximized the f-measure during training for OOB observations, some sensitivity and specificity values were taken from a suboptimal point in the ROC curve, leading to sensitivity values very close to or even equal to zero.

**eTable 5.** Prediction ability of machine learning for multisite pain incidence among adolescents. Results for the full set, balanced with SMOTE-NC.

| | | Prediction ability | | |
|---|---|---|---|---|
| **Multisite pain incidence** | **Cases/N** | **AUC (95 % CI)** | **Sensitivity** | **Specificity** |
| **All body sites** | | | | |
| Boys | 28/169 | 0.61 (0.49 to 0.72) | 0.07 (0.00 to 0.15) | 0.97 (0.94 to 1.00) |
| Girls | 65/230 | 0.69 (0.61 to 0.77) | 0.09 (0.01 to 0.17) | 0.96 (0.91 to 1.00) |
| **Musculoskeletal sites** | | | | |
| Boys | 18/172 | 0.60 (0.47 to 0.73) | 0.00[a] | 1.00[a] |
| Girls | 35/230 | 0.54 (0.45 to 0.63) | 0.03 (0.00 to 0.07) | 0.98 (0.96 to 1.00) |

Results are estimated from the validation set observations using the 10-fold cross-validation. [a]Due to automated threshold selection that maximized the f-measure during training for OOB observations, some sensitivity and specificity values were taken from a suboptimal point in the ROC curve, leading to sensitivity values very close to or even equal to zero.

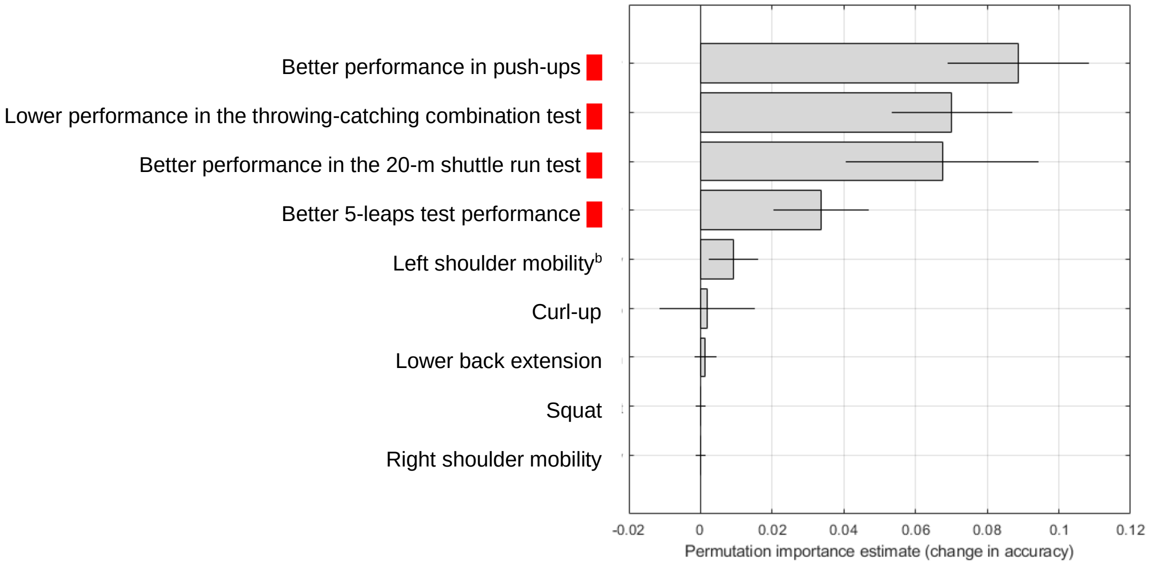


**eFigure 2.** Permutation importance estimates for boys in the *Move! set* for *all sites* (AUC 0.59). Red panel, risk factors; [b]Direction of the association not calculated for nominal variables

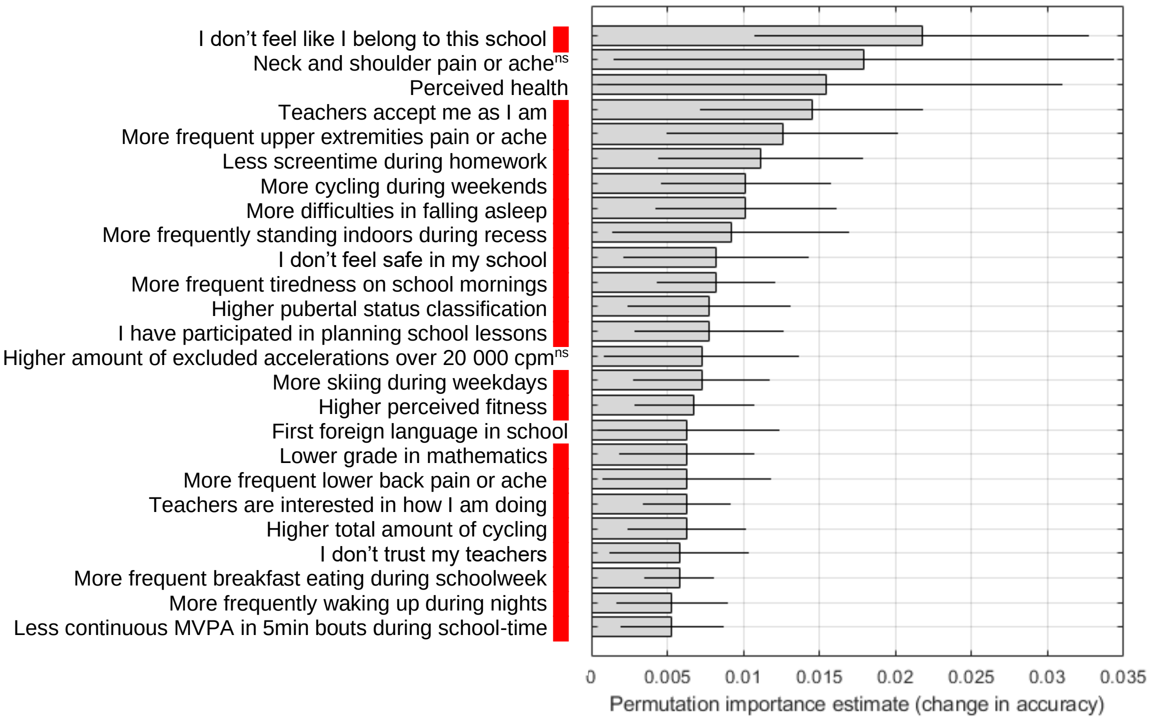


**eFigure 3.** Permutation importance estimates for girls in the *full set* for *all sites* (AUC 0.68). Only the top 25 predictors are presented. Red panel, risk factors; [ns]Not significant, variable significance calculated based on t-test in MATLAB, slightly differing from the manually calculated confidence intervals.

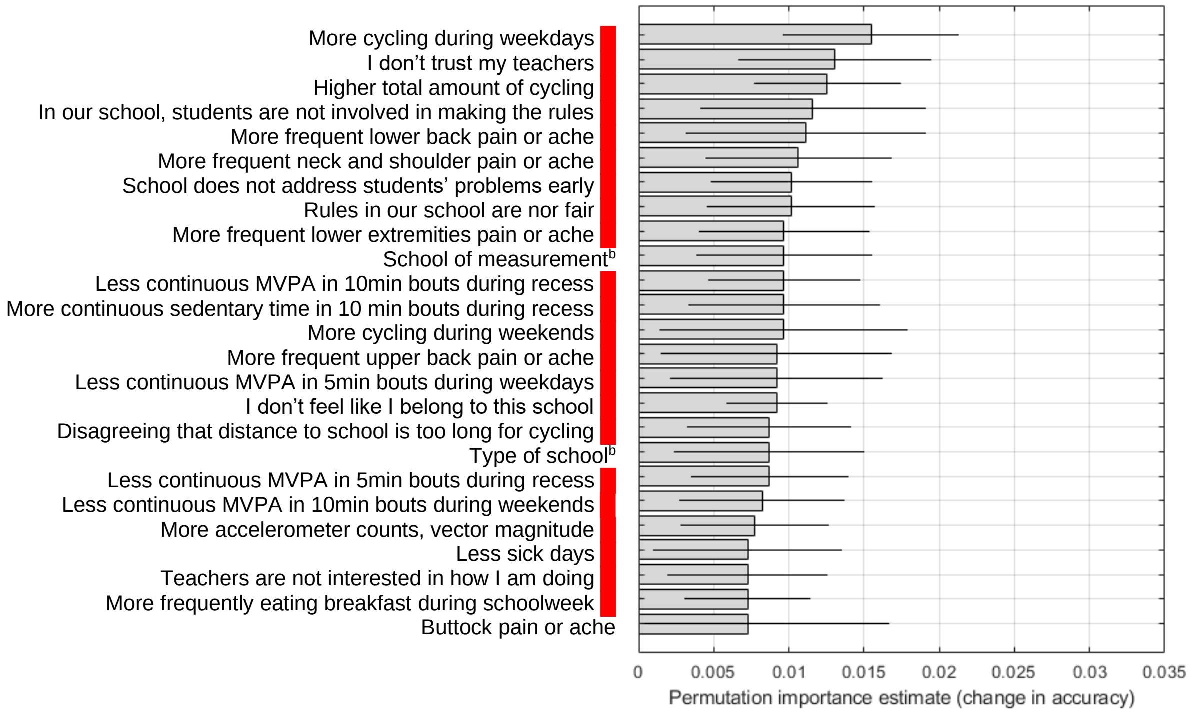


**eFigure 4.** Permutation importance estimates for girls in the *full set* for *musculoskeletal sites* (AUC 0.58). Only the top 25 predictors are presented. Red panel, risk factors; [b]Direction of the association not calculated for nominal variables

**Supplementary methods**

The training phase included procedures to optimize *pain* class prediction with RF and the training data. Bayesian optimization was employed to estimate the optimal hyperparameters (e.g. the method parameters that must be defined beforehand) for RF. The F-measure, which balances the precision and sensitivity of the classifier by computing their harmonic mean, was used as the optimization target for the RF out-of-bag samples. It is defined as

$$F = 2 \times \frac{precision \times sensitivity}{precision + sensitivity},$$

where

$$precision = \frac{TP}{TP + FP}$$

and

$$sensitivity = \frac{TP}{TP + FN}.$$

In the above equations, TP (true positives) refers to the number of correctly detected *pain* class cases. FP (false positives) is the number of cases incorrectly identified as belonging to the *pain* class and FN (false negatives) is the number of cases incorrectly identified as belonging to the *no pain* class. Since the Bayesian optimization aims to minimize the given objective, the final target for optimization was 1 – $F$.

The predicted probabilities of the RF model for the two classes were used to make receiver operating characteristic (ROC) curves, using OOB observations. During Bayesian optimization, the F-measure was taken from the point in the ROC curve that maximized the value. Afterwards, the best estimated hyperparameters were used when the RF model was trained again. The presented prediction results were then recorded using the OOB observations. Additionally, the threshold that maximized F-measure was recorded and later used when validating the results using the separate validation data portion in each fold. These separate results are presented only for the setup using SMOTE-NC, since the OOB estimates in RF are considered to be good for assessing prediction performance and generalizability.[1]

In addition to abovementioned equations, specificity, defined as

$$specificity = \frac{TN}{TN + FP},$$

was employed as a performance metric.

After training, a validation phase was implemented where the validity of the findings was tested against the left-out fold in 10-fold CV. During this phase, the measures used to estimate the prediction performance were AUC, sensitivity and specificity.

Additionally, accuracy, defined as

$$accuracy = \frac{TP + TN}{TP + FP + TN + FN}$$

was utilized in estimating the variable importances. Mean change in accuracy was used as the estimate for individual variable importance.

For handling the missing values in data, the original random forest method suggested two ways of imputing the missing values.[1] The TreeBagger implementation in MATLAB employs a surrogate decision split especially for handling the missing values in data. When the surrogate decision splits flag is set to “on”, a similar or correlated predictor value is used instead of the missing value.

Four RF hyperparameters in MATLAB's TreeBagger function were optimized:
1. *NumPredictorsToSample*: The number of variables to select at random for each decision split (range to search was from 1 to *total_number_of_variables_in_data-1*)
2. *MinLeafSize*: The minimum number of observations per tree leaf (range from 2 to 15).
3. *MaxNumSplits*: The maximum number of decision splits (range from 1 to 30).
4. *Surrogate*: Surrogate decision splits flag (options included on, off and all).

Static modified RF parameters included:
1. The number of trees in the forest was set to 500.
2. Nominal variables in the data were set as categorical variables (option *CategoricalPredictors*).
3. Algorithm used to select the best split predictor (option *PredictorSelection*) was set to *interaction-curvature*.

In addition, two static parameters were modified in MATLAB's Bayesian optimization (bayesopt) function:
1. *MaxObjectiveEvaluations* was set to 30, meaning that there are 30 iterations to search for optimal hyperparameters, after which the optimization was terminated.
2. *AcquisitionFunctionName* was set to expected-improvement-plus.

**References**
1. Breiman L. Random Forests. *Mach Learn*. 2001;45(1):5-32. doi:10.1023/A:1010933404324
